# Influence of socio-ecological factors on COVID-19 risk: a cross-sectional study based on 178 countries/regions worldwide

**DOI:** 10.1101/2020.04.23.20077545

**Authors:** Dai Su, Yingchun Chen, Kevin He, Tao Zhang, Min Tan, Yunfan Zhang, Xingyu Zhang

**Affiliations:** Department of Health Management, School of Medicine and Health Management, Tongji Medical College, Huazhong University of Science and Technology, Wuhan, China; Research Center for Rural Health Services, Hubei Province Key Research Institute of Humanities and Social Sciences, Wuhan, China; Department of Biostatistics, University of Michigan School of Public Health, Ann Arbor, United States; Department of Epidemiology and Health Statistics, West China School of Public Health and West China fourth Hospital, Sichuan University, Sichuan, China; Department of Systems, Populations, and Leadership, University of Michigan School of Nursing, Ann Arbor, United States

**Keywords:** socio-ecological factors, COVID-19 risk, cross-sectional study, 178 countries/regions worldwide

## Abstract

**Background:** The initial outbreak of COVID-19 caused by SARS-CoV-2 in China in 2019 has been severely tested in other countries worldwide. We aimed to describe the spatial distribution of the COVID-19 pandemic worldwide and assess the effects of various socio-ecological factors on COVID-19 risk.

**Methods:** We collected COVID-19 pandemic infection data and social-ecological data of 178 countries/regions worldwide from three database. We used spatial econometrics method to assess the global and local correlation of COVID-19 risk indicators for COVID-19. To estimate the adjusted incidence rate ratio (IRR), we modelled negative binomial regression analysis with spatial information and socio-ecological factors.

**Findings:** The study indicated that 37, 29 and 39 countries/regions were strongly opposite from the IR, CMR and DCI index “spatial autocorrelation hypothesis”, respectively. The IRs were significantly positively associated with GDP per capita, the use of at least basic sanitation services and social insurance program coverage, and were significantly negatively associated with the proportion of the population spending more than 25% of household consumption or income on out-of-pocket health care expenses and the poverty headcount ratio at the national poverty lines. The CMR was significantly positively associated with urban populations, GDP per capita and current health expenditure, and was significantly negatively associated with the number of hospital beds, number of nurses and midwives, and poverty headcount ratio at the national poverty lines. The DCI was significantly positively associated with urban populations, population density and researchers in R&D, and was significantly negatively associated with the number of hospital beds, number of nurses and midwives and poverty headcount ratio at the national poverty lines. We also found that climatic factors were not significantly associated with COVID-19 risk.

**Conclusion:** Countries/regions should pay more attention to controlling population flow, improving diagnosis and treatment capacity, and improving public welfare policies.

## 1. Introduction

The novel coronavirus disease (COVID-19) that has spread to more than one hundred countries and killed hundreds of thousands of people has officially been categorized as a pandemic by the World Health Organization. The initial outbreak of COVID-19 caused by SARS-CoV-2 in China in 2019 has been severely tested in other countries worldwide. As of April 6, 2020, COVID-19 had infected 1,345,048 patients in 184 countries/regions and caused 74,565 deaths, as countries worldwide responded to a human-to-human respiratory disease pandemic caused by COVID-19. Emerging infectious diseases (EIDs), such as SARS and COVID-19, pose a vast economic and public health burden worldwide [1,2].

COVID-19 not only seriously endangers people’s life safety and health but also greatly affects economic globalization. To address the challenges posed by COVID-19, the links among the transmission of COVID-19, socio-economic factors and climatic factors must be understood to suggest better strategies for predicting, preventing, coping with and mitigating the associated challenges. Simultaneously, given that the climate and socio-economic context are unlikely to change in the short term, it is easier to intervene accordingly [3]. The spread of many EIDs has been reported to be influenced by socio-ecological factors, including socio-economic and climate factors [1,2,4–7]. Previous studies have found that climatic conditions limit the geographical and seasonal distribution of EIDs, and weather affects the timing and intensity of outbreaks [8–12]. In addition, whereas climate patterns may control the potential global distribution of EIDs, the actual size and spatial scope of a region may be controlled by several non-climatic factors associated with transmission, including epidemiological, socio-economic and demographic factors [13–18]. However, research on the climatic and socio-economic drivers of COVID-19 transmission remains lacking, especially regarding the effects of socioeconomic factors and the total effects of socio-ecological factors. Ignoring important non-climatic factors or other confounding factors (such as urban development, economic growth, poverty, health, infrastructure, science and technology, social security and labor) would overestimate the effects of climate change. Therefore, studying the influence of socio-ecological factors on the transmission risk of EIDs is highly important.

For most EIDs, three elements are essential: an agent (or pathogen), a host (or vector) and the environment of transmission [19]. Appropriate climatic and weather conditions are necessary for the survival, reproduction, distribution and transmission of disease pathogens, vectors and hosts. Therefore, changes in climate or weather conditions may affect EIDs by affecting pathogens, vectors, hosts and their living environments [19–21]. Although many climate variables may influence the transmission of EIDs, some studies have shown that changes in the four main variables have the greatest effects on infected diseases with strong environmental components (temperature, precipitation, relative humidity (RH) and wind) [22–26]. In recent studies, although the severity of some cases of COVID-19 has mimicked that of SARS-CoV cases [27–30], the reproductive number (average R0=3.28) of COVID-19 is higher than that of SARS-CoV; therefore, considering the climate and environment may improve understanding of the pathogen’s vectorial capacity and basic reproduction number, and the risk of transmission of COVID-19 [31,32].

In recent decades, many rapid and pronounced changes in human social ecology have altered the likelihood of the emergence and spread of infectious diseases [33–35]. These changes include increases in population size and density; urbanization; persistent poverty (especially in the expansion of urban slums); the number and movement of political, economic and environmental refugees; differences in infrastructure and science and technology; and poor health awareness [36]. The socio-economic environment contributes significantly to the health of individuals as well as communities [37] and is the root cause of health and health equity. These socio-economic drivers have contributed to the shifting global ecology of vector transmission that enabled COVID-19 to emerge worldwide, by dangerously uniting the human hosts, vectors and pathogen. Socioeconomic changes interact with environmental changes in promoting EID spread and increase the harm of EIDs to humans.

The purpose of this study was to describe the spatial distribution of the COVID-19 pandemic worldwide, and assess the effects of different socio-ecological factors, including climate and socio-economic factors, on COVID-19 risk in 178 countries/regions worldwide, including incidence rate (IR), cumulative mortality rate (CMR) and daily cumulative index (DCI). In addition, this study analyzed intervention policies in different countries and regions to establish early warning and decision support systems and provide guidance for COVID-19 management in different countries/regions.

## 2. Methods

### 2.1. Concept model

According to previous research [38–41], we established the Potential Risk Assessment Framework for COVID-19 (Figure 1). The influence of global socio-ecological factors (climate and socio-economic factors) on the risk of COVID-19 can be tested by its influence on the following three disease components: agent (or pathogen), host (or vector) and the environment of transmission. A combination of natural and human influences led to the COVID-19 pandemic. We used three main variables to assess the potential risk of COVID-19: IR, CMR and DCI.

**Figure 1.**
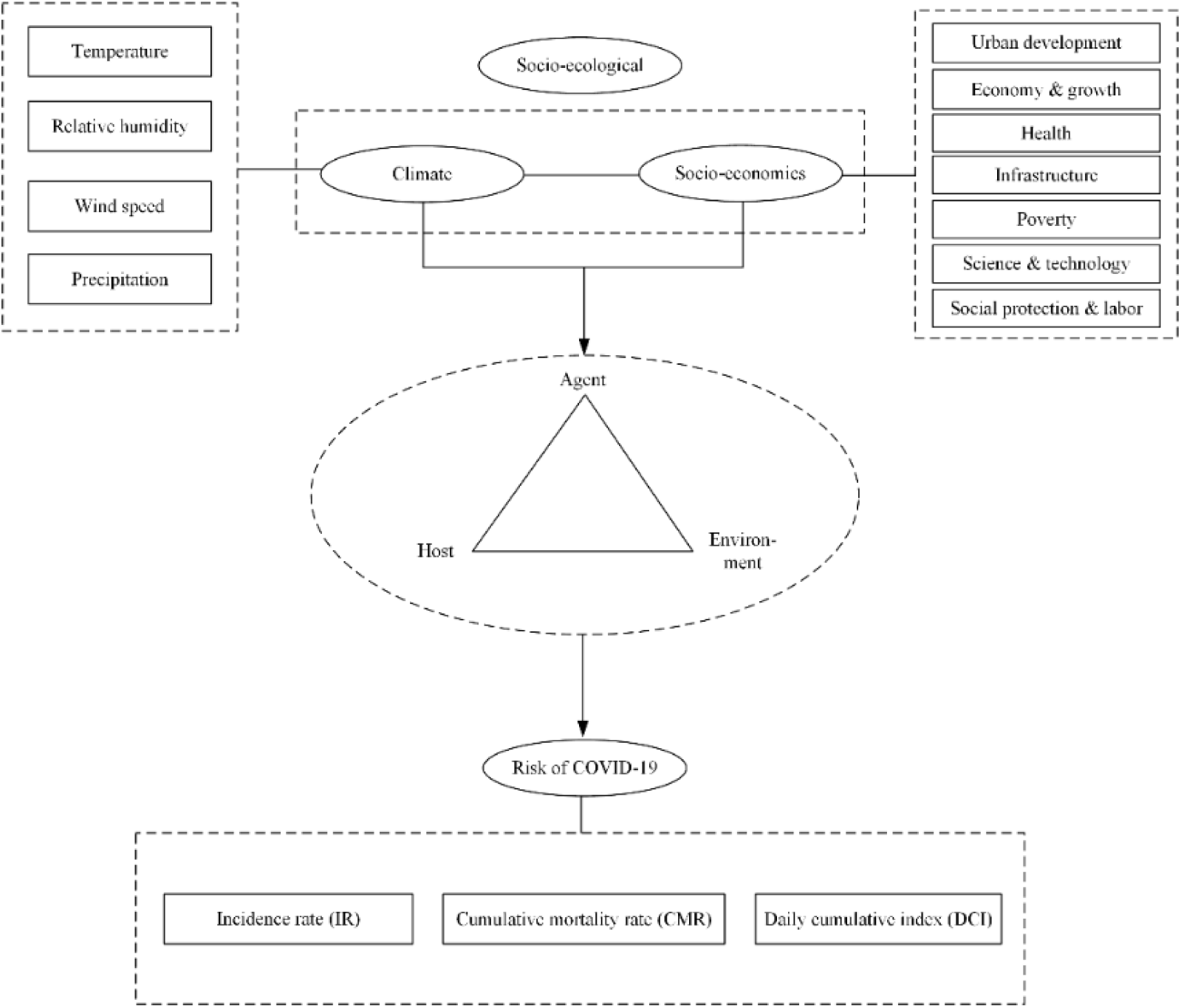
Climate, socio-economic conditions and COVID-19 transmission

### 2.2 Definitions of different cases for COVID-19

A confirmed case of COVID-19 infection was defined by laboratory confirmation of the virus causing COVID-19 infection, regardless of clinical signs and symptoms [42–44]. However, some reported case numbers from China have included people with symptoms of COVID-19 without laboratory confirmation. The definitions of COVID-19 related deaths differ across countries. In Italy, any death of a person with positive reverse transcriptase–polymerase chain reaction (RTPCR) testing for SARS-CoV-2 is considered COVID-19 related.

### 2.3 Data collection

#### 2.3.1 Outcome variables

A dashboard published and hosted by researchers at the Center for Systems Science and Engineering, Johns Hopkins University (JHU-CSSE) [45] shows the numbers and locations of confirmed COVID-19 cases, deaths and recoveries in all affected countries. All collected data on COVID-19 from the Johns Hopkins University are made freely available by the researchers through a GitHub repository. All manual updates (for countries and regions outside mainland China) are coordinated by a team at Johns Hopkins University. We extracted the global time series data of confirmed and recovered cases and deaths due to COVID-19 from the JHU-CSSE GitHub repository. The data were recorded from January 22, 2020 and were updated once daily around 23:59 (UTC). We selected the cross-sectional data from April 6, 2020. On the basis of the availability of the data, we extracted 178 countries from the database (excluding countries/regions without COVID-19 cases and some unmatched countries/regions, such as Taiwan, China). The first-level geographical unit of the dataset is the country/region, and the second-level geographical unit is the province/state. We uniformly selected first-level geographical units (countries/regions). In addition, we further classified the countries/regions in the data set according to UN geographical divisions and divided the countries with epidemic COVID-19 into 20 regions. As previously mentioned, for outcome variables, we selected IR, CMR and DCI as indicators to measure COVID-19 risk. The specific calculation process is shown in Table 1.

**Table 1.** The calculation process of outcome variables for COVID-19

| Outcome variables | Calculation process |
| --- | --- |
| Incidence rate (IR) | $IR = \frac{\text{Number of new cases of COVID-19 in a given time period}}{\text{Total population at risk during the follow-up period}} \times 1,000,000$ |
| Cumulative mortality rate (CMR) | $CMR = \frac{\text{number of COVID-19 deaths in a given time period}}{\text{total population during a given time period}} \times 1,000,000$ |
| Daily cumulative index (DCI) | $DCI = \frac{\text{cumulative COVID-19 confirmed cases}}{\text{number of days between the first reported case until now}}$ |

IR was used to describe the distribution of COVID-19, explore the etiological factors, propose an etiological hypothesis, and evaluate the efficacy of detection and prevention measures. CMR reflects the total deaths due to COVID-19 and is an indicator of the risk of death from COVID-19. DCI mainly describes the growth rate of COVID-19 in different countries/regions and is a measure of the risk of disease transmission. The World Health Organization, on March 11, 2020, declared the COVID-19 outbreak a global pandemic, thus indicating that COVID-19 had broadly spread worldwide. Therefore, when we considered IR, CMR and DCI in different countries/regions, these measures reflected not only the rapid growth in the number of people infected with COVID-19 but also the detection level in the entire country/region, which was used to identify more people infected with COVID-19.

#### 2.3.2 Climate data

We obtained daily meteorological observation values from the Global Surface Summary of the Day (GSOD) via The Integrated Surface Hourly (ISH) dataset. The ISH dataset includes global data obtained from the USAF Climatology Center, which is located in the Federal Climate Complex with NCDC. GSOD comprises 12 daily averages computed from global hourly station data. Except in United States stations, 24-hour periods are based on UTC times. The latest daily summary data are normally available 1–2 days after the date-time of the observations used in the daily summaries. More than 9,000 stations’ data worldwide are typically available. Daily weather elements include mean values of temperature, dew point temperature, sea level pressure, station pressure, visibility, wind speed, maximum and minimum temperature, maximum sustained wind speed and maximum gust, precipitation amount, snow depth and weather indicators. However, we chose the climate data from April 6, 2020 and selected four variables from the GSOD dataset that significantly affected COVID-19 risk: (1) mean temperature (.1 Fahrenheit); (2) mean dew point (.1 Fahrenheit); (3) mean wind speed (.1 knots); and (4) precipitation amount (.01 inches). The reason for extracting the average dew point variable was to calculate the RH value by using this variable and the temperature variable. The temperature and dew point in Celsius were used to calculate the RH according to the temperature and dew point at each time point [46]:

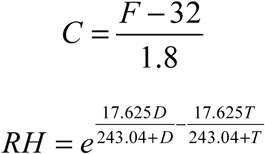

where *C* is the temperature in Celsius, *F* is the temperature in Celsius, *D* is the mean dew point for the day in Celsius, *T* is the mean temperature for the day in Celsius, and *e* is the base of the natural log.

#### 2.3.3 Socio-economic data

Indicators of socio-economic factors affecting the spread of COVID-19 were derived from the World Development Indicators dataset, the primary World Bank collection of development indicators, which compiles relevant, high-quality and internationally comparable statistics about global development and the fight against poverty. The database contains 1,600 time series indicators for 217 economies and more than 40 country groups, and data for many indicators cover a period of more than 50 years. As shown in Table 2, we selected 32 indicators affecting COVID-19 risk in seven dimensions in 178 countries/regions. The index value for 2019 was taken as the priority for each indicator. If the index was missing in 2019, the index value of the most recent year was selected as a substitute.

**Table 2.** Socioeconomic indicators influencing the spread of COVID-19

| Dimensions | Indicators | No. |
| --- | --- | --- |
| Urban Development | Urban population (% of total population) | (1) |
|  | Urban population growth (annual %) | (2) |
|  | Population density (people per sq. km of land area) | (3) |
| Economy & Growth | GDP per capita (current US\$) | (4) |
| Health | People using at least basic sanitation services (% of population) | (5) |
|  | Current health expenditure (% of GDP) | (6) |
| | Current health expenditure per capita (current US\$) | (7) |
|  | Death rate, crude (per 1,000 people) | (8) |
|  | Domestic private health expenditure (% of current health expenditure) | (9) |
| | Domestic private health expenditure per capita (current US\$) | (10) |
|  | Hospital beds (per 1,000 people) | (11) |
|  | Life expectancy at birth, total (years) | (12) |
|  | Maternal mortality ratio (national estimate, per 100,000 live births) | (13) |
|  | Mortality rate, infant (per 1,000 live births) | (14) |
|  | Net migration | (15) |
|  | Nurses and midwives (per 1,000 people) | (16) |
|  | People with basic handwashing facilities including soap and water (% of population) | (17) |
|  | Physicians (per 1,000 people) | (18) |
|  | Population aged 65 and above (% of total population) | (19) |
|  | Population growth (annual %) | (20) |
|  | Proportion of population spending more than 10% of household consumption or income on out-of-pocket health care expenses (%) | (21) |
|  | Proportion of population spending more than 25% of household consumption or income on out-of-pocket health care expenses (%) | (22) |
| Infrastructure | Railways, passengers carried (million passenger-km) | (23) |
| Poverty | Poverty headcount ratio at national poverty lines (% of population) | (24) |
| Science & Technology | Researchers in R&D (per million people) | (25) |
|  | Technicians in R&D (per million people) | (26) |
| Social Protection & Labor | Coverage of social insurance programs (% of population) | (27) |
|  | Unemployment, total (% of total labor force) (national estimate) | (28) |

### 2.4 Statistical analysis

#### 2.4.1. Spatial econometrics method

First, we used Moran’s I to measure the global correlation of COVID-19 risk indicators [47]. Global Moran’s I is a measure of global spatial autocorrelation, and the value of Moran’s I usually ranges from −1 to +1. Values significantly below -1/(N-1) indicate negative spatial autocorrelation, and values significantly above -1/(N-1) indicate positive spatial autocorrelation. If significant global spatial autocorrelation was found, we then used local indicators of spatial autocorrelation (LISA) to evaluate the locations of COVID-19 clusters. The meaning of local Moran’s Ii is similar to that of global Moran’s I. A positive Ii indicates that the high (or low) value of region i is surrounded by the surrounding high (or low) value; A negative Ii indicates that the high (or low) value of region i is surrounded by the surrounding low (or high) value. The general models are described in Eq. 1–2.

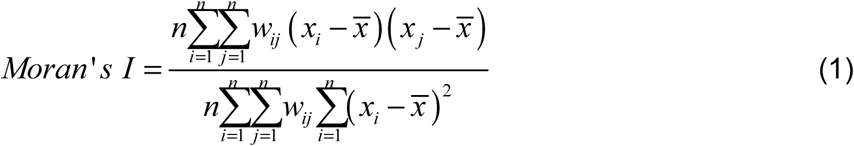

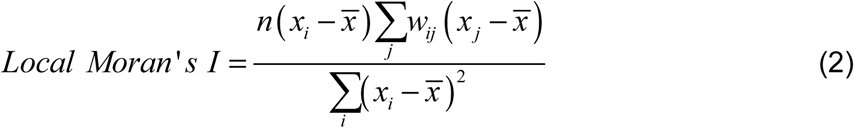

where *n* is the number of spatial units indexed by *i* and *j*, *x* is the variable of social ecology factors, 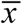 is the mean of *X*, and *w_ij_* is a matrix of spatial weights with zeroes on a diagonal (i.e., *w_ii_*=0).

Second, to better approximate the real infectious disease spatial spread process, we fit a one-order spatial autoregressive regression model comprising spatial lags. Because we believed that COVID-19 risk transmission in a certain country/region would be different for neighboring countries/regions, we sought to reflect this difference in the model. A one-order spatial autoregressive process takes the form (Eq. 3) [48]:

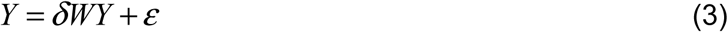

where *δ* is the spatial autoregressive coefficient, *W* is the *i*, *j*-th element of the exogenous,non-negative *N*×*N* spatial weight matrix with zero diagonal elements that describes the arrangement of the spatial units in the countries/regions, and *ε* is i.i.d. innovations with zero mean and finite variance *σ*^2^. For simplicity, in this paper, we assumed that the spatial weight matrix *W* was non-standardized and also used a queen spatial weight matrix.

#### 2.4.2. Processing of missing values

Before negative binominal regression, the k-nearest neighbors (k-NN) approach was used to impute missing data for some socio-ecological variables. For a given patient with missing values, the k-NN method identifies the k-nearest countries/regions on the basis of Euclidean distance. Using these countries/regions, we then replaced missing values by using a majority vote for discrete variables and weighted means for continuous features. One advantage of using this method is that missing values in all features are imputed simultaneously without the need to treat features individually.

#### 2.4.3. Negative binomial regression

First, we established the correlation matrix of socio-economic factors to check for multicollinearity. If there was a strong correlation (> 0.8) among socio-economic factors, then we removed the factor with strong correlation with other variables. Then, the incidence rate ratio (IRR) of each socio-ecological factor was calculated with single factor negative binomial regression analysis, that is, the effect of each socio-ecological factor on COVID-19 risk by changing the average COVID-19 risk value by a specific unit quantity. The spatial autoregressive models comprising spatial lags, which were a weighted average of observations on the diseases over neighboring units, were input into the model to adjust for spatial variation in COVID-19 risk. Modeled values of climate factors were centered on the mean values for each station in every country/region [49]. The factors with P < 0.05 were included in the multi-factor negative binomial regression analysis with spatial information to calculate the adjusted IRR (aIRR). The general model is described in Eq. 4.

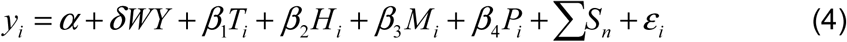

where *y_i_* denotes the daily counts of COVID-19 risk indicators in county/region *i*; *WY* represents spatial lags, and *W*; is the spatial weight; *S_n_* represents socio-economic factors (all variables are in Table 2); *T_t_* is the mean temperature in county/region *i*; *H_t_* is the RH in county/region *i*; *M_t_* is the wind speed in county/region *i*; *P_t_* is the precipitation amount in county/region *i*; and *ε_i_* is a random intercept.

Sensitivity analyses with maximum and minimum temperatures instead of average temperatures were also conducted with the same procedures, in which we used the same non-informative priors for the minimum and maximum temperatures [49, 50]. All statistical analyses were performed in Stata statistical software Version 15, and p-values were two-tailed, with statistical significance set at.05. ArcMap 10.7 and Geoda software were used to process basic geographic information. Data visualization mainly used RStudio software Version 1.2.5033.

## 3. Results

### 3.1 Characteristics of 178 countries/regions with reported cases of COVID-19

As of April 6, 2020, a total of 178 countries/regions worldwide had reported data and were included in this study (Table S1). The three countries/regions with the highest IR worldwide were Andorra (Southern Europe, IR=313.80), Iceland (Northern Europe, IR=215.90) and Gibraltar (United Kingdom) (Southern Europe, IR=178.52). The three countries/regions with the highest CMR worldwide were San Marino (Southern Europe, CMR=947.17), Spain (Southern Europe, IR=285.53) and Italy (Southern Europe, IR=273.42). The three countries/regions with the highest DCI worldwide were the United States (North America, DCI=4823.87), Spain (Southern Europe, DCI=2070.83) and Italy (Southern Europe, DCI=1978.31).

### 3.2 Spatial clustering evaluation for COVID-19

#### 3.2.1 Test results for global spatial correlation

The number and distribution of first-order neighbors in different countries/regions are shown in Figure 2. The number of neighboring countries/regions was mainly concentrated in 0-6, accounting for 87.08% of the total number of neighboring countries. Among them, China and Russia had the largest number of neighboring countries/regions.

**Figure 2.**
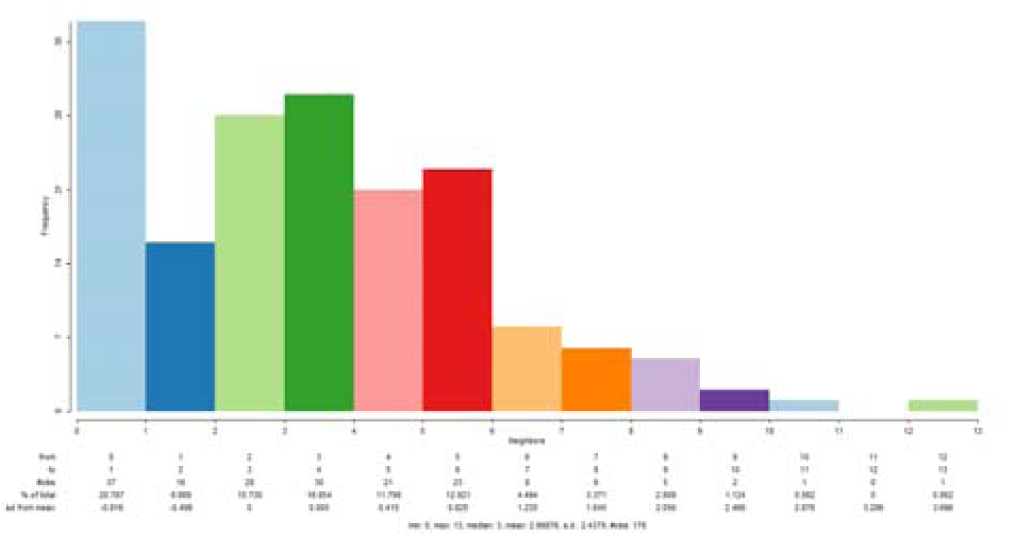
Number of first-order neighbors in different countries/regions

Table S2 shows the global spatial Moran’s I indexes of the IR, CMR and DCI of 178 countries/regions worldwide according to the one-order spatial contiguity matrix. The Moran’s I indexes of IR, CMR and DCI were all positive, and all index values were significant at the level of 1%, thus indicating that the IR, CMR and DCI of 178 countries/regions worldwide had strong spatial aggregation effects. Meanwhile, the Moran’s I index of different indicators showed significant differences, thus indicating to some extent that IR, CMR and DCI have different aggregation effects in different countries/regions.

#### 3.2.2 Test results for local spatial correlation

A total of 37 countries/regions were strongly opposite from the IR index “spatial autocorrelation hypothesis,” including 11 countries/regions with high–high patterns, mainly concentrated in Western Europe, southern Europe and Canada; 24 countries/regions with low–low patterns, mainly concentrated in parts of Africa, parts of Asia (China, India, Laos); Cuba with a low–high pattern; and Djibouti with a high–low pattern. Simultaneously, 29 countries/regions were strongly opposite from the CMR index “spatial autocorrelation hypothesis,” which was suitable for 10 countries with high–high patterns, mainly in Western Europe and southern Europe (e.g., France, Italy and Spain); 16 countries/regions with low–low patterns, mainly concentrated in parts of Africa and China; and three countries with low–high patterns, including Morocco and Slovenia. In addition, 39 countries/regions strongly did not support the hypothesis of “no spatial autocorrelation” of the DCI index, among which six countries/regions had high–high patterns (Canada, France, Portugal, Belgium, the Netherlands, and Switzerland), and 22 countries/regions had low–low patterns, mainly in parts of Africa and Honduras. Eleven countries were in the low–high pattern category, including Mexico, Cuba, Morocco, Denmark and Luxembourg, etc. The above results are consistent with the global spatial autocorrelation test results, thus indicating that IR, CMR and DCI indicators in some countries/regions may be affected by the COVID-19 epidemic in neighboring countries/regions and may show clear geographical characteristics.

To directly reflect the local spatial characteristics of IR, CMR and DCI, LISA scatter diagrams of the three indexes are shown in Figure 3. Most of the three indicators fell into the third quadrant (low–low), but the countries/regions whose IR and DCI index fell into the first quadrant (high– high) and the second quadrant (low–high) had indicator values exceeding the CMR. Thus, among the 178 countries/regions worldwide, the countries/regions with low IR, CMR and DCI indicators showed a spatial agglomeration effect, as did the countries/regions with high IR, CMR and DCI indicators. In addition, some neighboring countries/regions showed some differences in IR, CMR and DCI (high–low and low–high).

**Figure 3.**
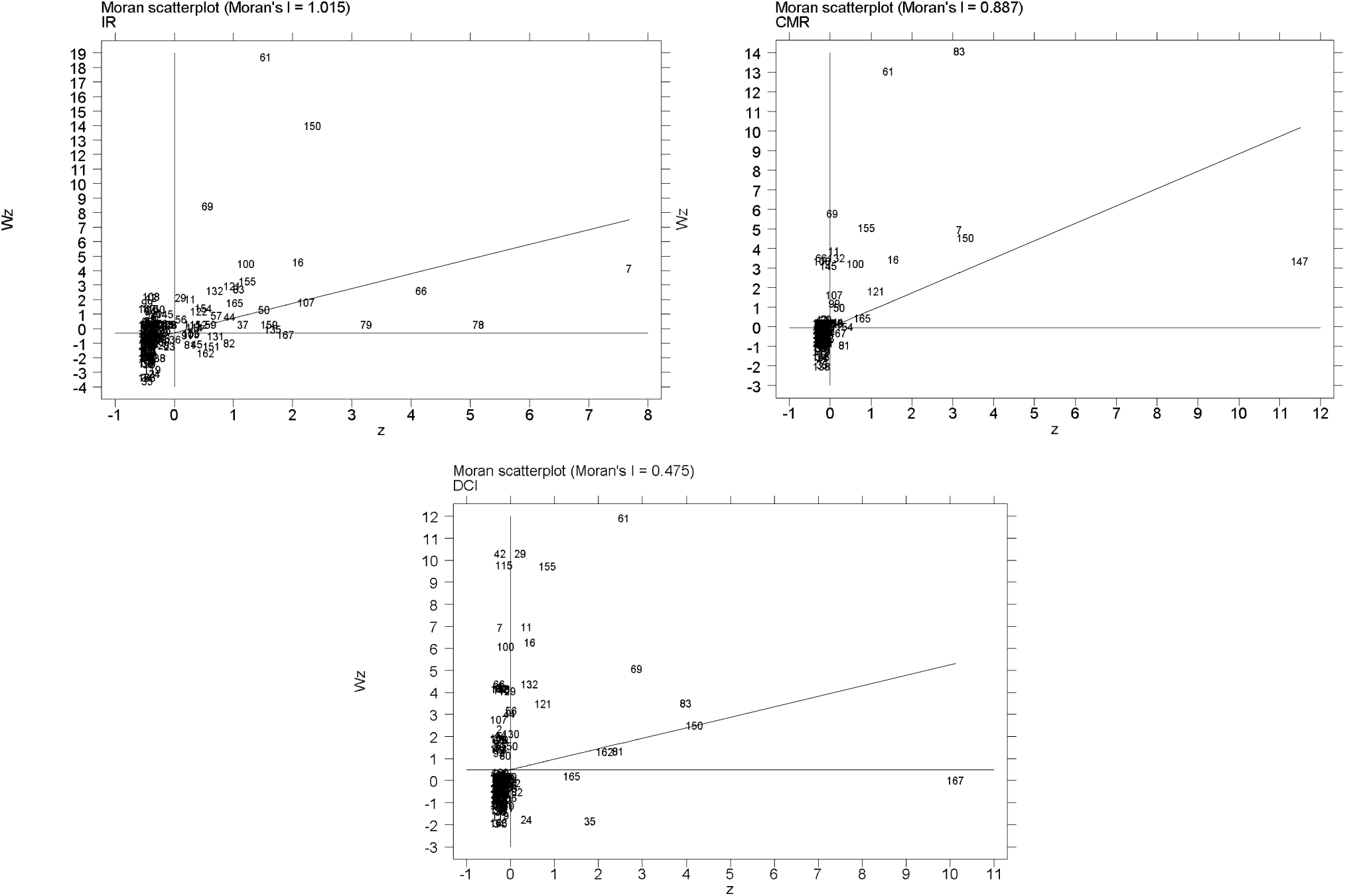
LISA scatter diagram of IR, CMR and DCI indexes for COVID-19 in 178 countries/regions. (z is the value of the variable, and Wz is the local Moran’s Ii value of the variable.)

### 3.3 Analysis of the influence of socio-ecological factors on COVID-19 risk

#### 3.3.1 Correlation analysis of socio-economic factors

To eliminate the influence of the collinearity between the socio-economic indicators on the estimation effect of the model, we established a correlation matrix of the socio-economic indicators (Table S3). The indexes with strong correlation (> 0.8) were screened, and one of the effective indexes was reserved for model analysis. We excluded eight socio-economic indicators in Table 2, numbered 7 (current health expenditure per capita), 12 (total life expectancy at birth), 13 (maternal mortality ratio), 14 (infant mortality rate), 17 (access to basic handwashing facilities including soap and water), 20 (population growth), 21 (proportion of the population spending more than 10% of household consumption or income on out-of-pocket health care expenditure) and 26 (technicians in R&D), and we retained 20 socio-economic indicators.

#### 3.3.2 Negative binomial regression analysis of socio-ecological factors on COVID-19 risk

We analyzed the effects of socio-ecological factors on COVID-19 risk in 178 countries. The results of single-factor and multi-factor negative binomial regression analysis are shown in Table 4.

**Table 3.** Characteristics of UN geographical divisions with reported cases of COVID-19 as of 6 April 2020

| UN geographical divisions | No. of Countries/regions | IR† | CMR‡ | DCI§ | Total population | Total days since first reported case |
| --- | --- | --- | --- | --- | --- | --- |
| Africa |  |  |  |  |  |  |
| Eastern Africa | 17 | 0.18 | 0.04 | 2.85 | 451173502 | 325 |
| Middle Africa | 8 | 0.12 | 0.21 | 4.96 | 168910830 | 189 |
| Northern Africa | 5 | 1.92 | 1.85 | 25.03 | 194924933 | 179 |
| Southern Africa | 3 | 0.50 | 0.21 | 26.28 | 62482003 | 65 |
| Western Africa | 13 | 0.35 | 0.16 | 4.93 | 343169384 | 299 |
| Asia |  |  |  |  |  |  |
| Central Asia | 3 | 4.55 | 0.21 | 19.63 | 57547699 | 68 |
| East Asia | 4 | 0.40 | 2.29 | 355.28 | 1574064564 | 273 |
| Southeast Asia | 9 | 1.47 | 0.78 | 26.75 | 600191804 | 525 |
| Southern Asia | 9 | 2.18 | 2.09 | 159.97 | 1896189013 | 436 |
| Western Asia | 14 | 11.32 | 1.67 | 36.21 | 138585525 | 586 |
| Europe |  |  |  |  |  |  |
| Eastern Europe | 10 | 6.91 | 1.85 | 64.50 | 292450026 | 379 |
| Northern Europe | 12 | 51.86 | 60.72 | 153.93 | 104678611 | 528 |
| Southern Europe | 14 | 64.24 | 204.80 | 535.26 | 148744007 | 541 |
| Western Asia | 1 | 38.24 | 7.88 | 1119.15 | 82319724 | 27 |
| Western Europe | 9 | 57.69 | 77.55 | 628.85 | 196616151 | 442 |
| North America |  |  |  |  |  |  |
| Caribbean | 16 | 3.35 | 2.99 | 7.62 | 38841019 | 338 |
| Central America | 8 | 2.86 | 1.03 | 24.87 | 175471759 | 203 |
| North America | 4 | 83.30 | 30.53 | 2027.60 | 364346283 | 189 |
| Oceania |  |  |  |  |  |  |
| Australia and New Zealand | 2 | 5.92 | 1.37 | 62.19 | 29877869 | 111 |
| Melanesia | 5 | 0.49 | 0.00 | 1.92 | 22465856 | 106 |
| South America | 12 | 4.74 | 2.39 | 74.46 | 423398995 | 367 |

†IR, incidence rate (per 10 million people).
‡CMR, cumulative mortality rate (per 10 million people).
§DCI, daily cumulative index (%)

**Table 4.** The results of single-factor and multi-factor negative binomial regression analysis for COVID-19 in 178 countries/regions

| Indicators | Incidence rate (IR) |  |  |  |  |  |  |  | Cumulative mortality rate (CMR) |  |  |  |  |  |  |  | Daily cumulative index (DCI) |  |  |  |  |  |  |  |
| --- | --- | --- | --- | --- | --- | --- | --- | --- | --- | --- | --- | --- | --- | --- | --- | --- | --- | --- | --- | --- | --- | --- | --- | --- |
|  | IRR* | 95%CI†<br>(Lower) | 95%CI<br>(Upper) | p-value | aIRR‡ | 95%CI<br>(Lower) | 95%CI<br>(Upper) | p-value | IRR | 95%CI<br>(Lower) | 95%CI<br>(Upper) | p-value | aIRR | 95%CI<br>(Lower) | 95%CI<br>(Upper) | p-value | IRR | 95%CI<br>(Lower) | 95%CI<br>(Upper) | p-value | aIRR | 95%CI<br>(Lower) | 95%CI<br>(Upper) | p-value |
| **Urban Development** |  |  |  |  |  |  |  |  |  |  |  |  |  |  |  |  |  |  |  |  |  |  |  |  |
| Urban population (% of total population) | 1.048 | 0.992 | 1.106 | 0.096 |  |  |  |  | 1.055 | 1.027 | 1.083 | <0.001 | 1.027 | 1.010 | 1.044 | 0.001 | 1.073 | 1.055 | 1.091 | <0.001 | 1.022 | 1.010 | 1.034 | <0.001 |
| Urban population growth (annual %) | 1.053 | 1.039 | 1.066 | <0.001 | 0.866 | 0.670 | 1.120 | 0.273 | 0.287 | 0.203 | 0.405 | <0.001 | 0.751 | 0.507 | 1.112 | 0.153 | 0.407 | 0.312 | 0.530 | <0.001 | 0.993 | 0.734 | 1.343 | 0.963 |
| Population density (people per sq. km of land area) | 1.000 | 1.000 | 1.000 | 0.179 |  |  |  |  | 1.000 | 1.000 | 1.000 | 0.633 |  |  |  |  | 1.000 | 1.000 | 1.000 | <0.001 | 1.000 | 1.000 | 1.000 | 0.020 |
| **Economy & Growth** |  |  |  |  |  |  |  |  |  |  |  |  |  |  |  |  |  |  |  |  |  |  |  |  |
| GDP per capita (current 1,000 US\$) | 1.054 | 1.038 | 1.071 | <0.001 | 1.029 | 1.013 | 1.045 | <0.001 | 1.086 | 1.056 | 1.117 | <0.001 | 1.031 | 1.021 | 1.041 | <0.001 | 1.045 | 1.019 | 1.072 | 0.001 | 0.998 | 0.987 | 1.009 | 0.696 |
| **Health** |  |  |  |  |  |  |  |  |  |  |  |  |  |  |  |  |  |  |  |  |  |  |  |  |
| People using at least basic sanitation services (% of population) | 1.063 | 1.048 | 1.078 | <0.001 | 1.022 | 1.005 | 1.039 | 0.010 | 1.088 | 1.065 | 1.111 | <0.001 | 1.001 | 0.985 | 1.018 | 0.897 | 1.056 | 1.045 | 1.066 | <0.001 | 1.011 | 0.998 | 1.025 | 0.102 |
| Current health expenditure (% of GDP) | 1.186 | 1.045 | 1.345 | 0.008 | 1.088 | 0.956 | 1.238 | 0.203 | 1.501 | 1.245 | 1.809 | <0.001 | 1.211 | 1.040 | 1.410 | 0.013 | 1.416 | 1.216 | 1.648 | <0.001 | 1.080 | 0.968 | 1.206 | 0.169 |
| Death rate, crude (per 1,000 people) | 0.954 | 0.862 | 1.057 | 0.371 |  |  |  |  | 1.097 | 0.857 | 1.403 | 0.462 |  |  |  |  | 1.187 | 0.978 | 1.439 | 0.082 |  |  |  |  |
| Domestic private health expenditure (% of current health expenditure) | 0.955 | 0.933 | 0.978 | <0.001 | 0.988 | 0.972 | 1.004 | 0.135 | 0.937 | 0.906 | 0.969 | <0.001 | 0.986 | 0.970 | 1.003 | 0.102 | 0.951 | 0.933 | 0.970 | <0.001 | 1.010 | 0.994 | 1.027 | 0.207 |
| Domestic private health expenditure per capita (current US\$) | 1.003 | 1.002 | 1.003 | <0.001 | 1.001 | 1.000 | 1.001 | 0.119 | 1.005 | 1.003 | 1.008 | <0.001 | 1.001 | 1.000 | 1.001 | 0.104 | 1.003 | 1.001 | 1.004 | <0.001 | 1.000 | 1.000 | 1.001 | 0.438 |
| Hospital beds (per 1,000 people) | 1.246 | 1.034 | 1.502 | 0.021 | 0.906 | 0.790 | 1.039 | 0.156 | 1.798 | 1.067 | 3.030 | 0.028 | 0.799 | 0.696 | 0.916 | 0.001 | 1.302 | 1.022 | 1.659 | 0.033 | 0.731 | 0.641 | 0.833 | <0.001 |
| Net migration | 1.000 | 1.000 | 1.000 | <0.001 | 1.000 | 1.000 | 1.000 | 0.954 | 1.000 | 1.000 | 1.000 | 0.001 | 1.000 | 1.000 | 1.000 | 0.530 | 1.000 | 1.000 | 1.000 | 0.005 | 1.000 | 1.000 | 1.000 | 0.103 |
| Nurses and midwives (per 1,000 people) | 1.239 | 1.149 | 1.337 | <0.001 | 0.901 | 0.813 | 1.002 | 0.052 | 1.499 | 1.268 | 1.773 | <0.001 | 0.837 | 0.749 | 0.936 | 0.002 | 1.290 | 1.111 | 1.498 | 0.001 | 0.904 | 0.820 | 0.997 | 0.044 |
| Physicians (per 1,000 people) | 1.938 | 1.393 | 2.698 | <0.001 | 1.005 | 0.750 | 1.347 | 0.972 | 3.389 | 2.652 | 4.331 | <0.001 | 1.725 | 1.191 | 2.498 | 0.004 | 2.600 | 1.739 | 3.886 | <0.001 | 0.951 | 0.758 | 1.192 | 0.661 |
| Population ages 65 and above (% of total population) | 1.111 | 1.054 | 1.171 | <0.001 | 0.981 | 0.913 | 1.055 | 0.609 | 1.235 | 1.151 | 1.326 | <0.001 | 1.066 | 0.968 | 1.173 | 0.192 | 1.233 | 1.144 | 1.328 | <0.001 | 1.062 | 0.988 | 1.141 | 0.102 |
| Proportion of population spending more than 25% of household consumption or income on out-of-pocket health care expenditure (%) | 0.802 | 0.675 | 0.953 | 0.012 | 0.846 | 0.750 | 0.955 | 0.007 | 0.794 | 0.626 | 1.007 | 0.057 |  |  |  |  | 0.898 | 0.715 | 1.129 | 0.357 |  |  |  |  |
| **Infrastructure** |  |  |  |  |  |  |  |  |  |  |  |  |  |  |  |  |  |  |  |  |  |  |  |  |
| Railways, passengers carried (million passenger-km) | 1.000 | 1.000 | 1.000 | <0.001 | 1.000 | 1.000 | 1.000 | 0.323 | 1.000 | 1.000 | 1.000 | 0.679 |  |  |  |  | 1.000 | 1.000 | 1.000 | 0.498 |  |  |  |  |
| **Poverty** |  |  |  |  |  |  |  |  |  |  |  |  |  |  |  |  |  |  |  |  |  |  |  |  |
| Poverty headcount ratio at national poverty lines (% of population) | 0.924 | 0.900 | 0.949 | <0.001 | 0.970 | 0.948 | 0.993 | 0.011 | 0.894 | 0.872 | 0.916 | <0.001 | 0.960 | 0.940 | 0.982 | <0.001 | 0.908 | 0.889 | 0.926 | <0.001 | 0.963 | 0.945 | 0.982 | <0.001 |
| **Science & Technology** |  |  |  |  |  |  |  |  |  |  |  |  |  |  |  |  |  |  |  |  |  |  |  |  |
| Researchers in R&D (per million people) | 1.000 | 1.000 | 1.001 | <0.001 | 1.000 | 1.000 | 1.000 | 0.159 | 1.000 | 1.000 | 1.001 | 0.034 | 1.000 | 1.000 | 1.000 | 0.115 | 1.001 | 1.001 | 1.001 | <0.001 | 1.000 | 1.000 | 1.001 | 0.012 |
| **Social Protection & Labor** |  |  |  |  |  |  |  |  |  |  |  |  |  |  |  |  |  |  |  |  |  |  |  |  |
| Coverage of social insurance programs (% of population) | 1.060 | 1.041 | 1.079 | <0.001 | 1.047 | 1.009 | 1.086 | 0.014 | 1.036 | 0.992 | 1.082 | 0.110 |  |  |  |  | 1.091 | 1.072 | 1.110 | <0.001 | 1.015 | 0.991 | 1.039 | 0.214 |
| Unemployment, total (% of total labor force) (national estimate) | 0.902 | 0.828 | 0.982 | 0.018 | 0.956 | 0.909 | 1.004 | 0.072 | 1.002 | 0.861 | 1.166 | 0.984 |  |  |  |  | 0.986 | 0.866 | 1.122 | 0.828 |  |  |  |  |
| **Climate** |  |  |  |  |  |  |  |  |  |  |  |  |  |  |  |  |  |  |  |  |  |  |  |  |
| Mean temperature (Celsius) | 0.934 | 0.897 | 0.972 | 0.001 | 1.039 | 0.992 | 1.088 | 0.105 | 0.975 | 0.887 | 1.071 | 0.592 |  |  |  |  | 0.834 | 0.800 | 0.869 | <0.001 | 0.978 | 0.939 | 1.018 | 0.280 |
| Relative humidity (%) | 1.011 | 0.994 | 1.027 | 0.204 |  |  |  |  | 1.025 | 0.995 | 1.056 | 0.108 |  |  |  |  | 0.990 | 0.960 | 1.022 | 0.538 |  |  |  |  |
| Mean wind speed (.1 knots); | 1.210 | 1.111 | 1.317 | <0.001 | 1.040 | 0.957 | 1.130 | 0.358 | 1.155 | 0.951 | 1.403 | 0.146 |  |  |  |  | 0.958 | 0.836 | 1.097 | 0.533 |  |  |  |  |
| Precipitation amount (.01 inches). | 0.082 | 0.016 | 0.003 | 0.003 | 0.477 | 0.126 | 1.808 | 0.276 | 0.019 | 0.001 | 0.377 | 0.009 | 0.425 | 0.123 | 1.471 | 0.177 | 0.204 | 0.005 | 8.409 | 0.402 |  |  |  |  |

\*IRR, incidence rate ratio.
†CI is short for confidence interval.
‡aIRR, adjusted incidence rate ratio.

The IR was significantly positively associated with GDP per capita (aIRR=1.029, 95%CI: 1.013–1.045), use of at least basic sanitation services (aIRR=1.022, 95%CI: 1.005–1.039) and coverage of social insurance programs (aIRR=1.047, 95%CI: 1.009–1.086), and was significantly negatively associated with the proportion of the population spending more than 25% of household consumption or income on out-of-pocket health care expenses (aIRR=0.846, 95%CI: 0.750–0.955) and the poverty headcount ratio at national poverty lines (aIRR=0.970, 95%CI: 0.948–0.993).

The CMR was significantly positively associated with urban populations (aIRR=1.027, 95%CI: 1.010–1.044), GDP per capita (aIRR=1.031, 95%CI: 1.021–1.041) and current health expenditure (aIRR=1.211, 95%CI: 1.040–1.410), and was significantly negatively associated with the number of hospital beds (aIRR=0.799, 95%CI: 0.696–0.916), number of nurses and midwives (aIRR=0.837, 95%CI: 0.749–0.936) and poverty headcount ratio at the national poverty lines (aIRR=0.960, 95%CI: 0.940–0.982).

The DCI was significantly positively associated with urban populations (aIRR=1.021, 95%CI: 1.009–1.034), population density (aIRR=1.000, 95%CI: 1.000–1.000) and researchers in R&D (aIRR=1.000, 95%CI: 1.000–1.001), and was significantly negatively associated with the number of hospital beds (aIRR=0.731, 95%CI: 0.641–0.833), number of nurses and midwives (aIRR=0.904, 95%CI: 0.820–0.997) and poverty headcount ratio at the national poverty lines (aIRR=0.963, 95%CI: 0.945–0.982).

The results of the sensitivity analysis are reported in Table S4 and Table S5: we used the maximum and minimum temperatures instead of the average temperature, and then incorporated the two climate factors into the single-factor and multi-factor negative binomial regression. The results showed that the significance of different socio-ecological factors was essentially consistent. We found that only the variable of poverty headcount ratio at the national poverty lines (percentage of population) became significant after sensitivity analysis on IR, thus indicating that the analysis results were relatively reliable.

## 4. Discussion

By evaluating the spatial aggregation characteristics of three indicators—IR, CMR and DCI—on COVID-19 risk in 178 countries, Western Europe, Southern Europe, East Asia and some African countries, we found that all showed relatively large spatial correlations, thus indicating that COVID-19 broadly affects these countries/regions.

Because COVID-19 is highly contagious, after an outbreak occurs in a country/region, the virus tends to spread rapidly in surrounding countries. Italy was the first country in Europe to have a large outbreak of COVID-19, but the Italian health system adopted several control measures, such as timely intervention and containment measures brought about by decentralization, flexible financing mechanisms, private and public sector partnerships, and human resources mobilization, so that the IR and DCI could be effectively controlled [51]. However, because some European countries did not perform effective prevention and control measures, such as blockading countries or cities, in early stages of the outbreak, the epidemic gradually broke out in countries including France, Germany, Spain and Portugal. In North America, the development of COVID-19 presents progressive characteristics (high–high and low–high mode), and Canada is also significantly affect clearly the United States by the COVID–19 outbreak, but the DCI in Mexico and other countries/regions in Central America remained relatively low. The United States also recently closed its border with Canada and Mexico and decreased the flow of people across the border. Because, blockading and quarantining provide very good protection, taking these measures in countries or cities is very important to decrease the risk of COVID-19 multinational spread. The potential transmission of COVID-19 in South America must not be ignored.

Second, both IR and CMR presented low–low patterns in China, thus indicating that China and some neighboring countries (such as South Korea and Singapore) have effectively reduced the risk to neighboring countries by implementing strong prevention and control measures against COVID-19 transmission and have also acquired valuable experience useful to other countries in fighting COVID-19 virus [52–56]. However, notably, the IR of COVID-19 in India presents a low– low model, thus indicating that India is at low risk of COVID-19 transmission from surrounding countries, and consequently has a low DCI. However, as the world’s second most populous country, India may have a high risk of COVID-19 transmission because of inadequate medical conditions and detection levels. Simultaneously, China, South Korea and other countries must strengthen screening of imported cases from other countries, reduce social contact among travelers and prevent the possible secondary transmission of COVID-19 [52,57,58].

Third, most countries/regions in Africa remain in a low–low mode, but the short distance and frequent contacts between Western and Southern Europe and North Africa may place North Africa at high risk of COVID-19 spread; Morocco currently has a low–high mode representing an early warning, and COVID-19 viruses must further be prevented from entering other parts of Africa. Although African countries took measures to prevent the Ebola outbreak in 2014, Africa remains one of the poorest countries worldwide, and it has a shortage of health resources to quickly control the outbreak. Studies have shown that the current spread of COVID-19 in West Africa urgently requires action to control the further spread of COVID-19 and improve the response capacity of affected countries in West Africa [59]. Although most parts of Africa are in the low–low mode, they may also face threats. Many COVID-19 cases may be undetected, thus potentially explaining the current low-level indicators (IR, CMR and DCI).

We found that, in terms of urban development, both CMR and DCI were significantly positive associated with the urban population (percentage home of total population), and people per sq. km of land area had significant positive effects on the DCI. Non-drug intervention measures have already been implemented, and if traffic restrictions, social isolation and family measures are not ensured, the increase in population density and urbanization may result in many problems, such as public traffic, rural population health inequities, poor housing conditions, inadequate freshwater supply, and poor sanitation and ventilation systems, thus accelerating the spread of the COVID-19 virus, in agreement with previous research [60,61]. The higher the urban population (percentage of total population), the faster the urbanization process of the country/region; consequently, aging and young people participating in social activities become more likely to aggravate the spread of the virus and increase the burden on the health system, in agreement with the results of one study [62]. In addition, studies have shown that, with urbanization, the risk of infection and the chances of survival after COVID-19 infection among older individuals with complications is greatly increased, thus resulting in a significant increase in the CMR in the country/region [63–64].

In terms of the economy and growth, we found that GDP per capita (current 1,000 US$) was significantly positive associated with the IR and CMR of COVID-19, possibly because the GDP per capita tends to reflect a country’s economic development level: with higher GDP per capita, governments can invest more in screening and treatment of patients with mild and severe cases of COVID-19. Consequently, with more confirmed cases and deaths, classification strategies can be considered for COVID-19 in low-income groups. Especially in economically underdeveloped areas such as Africa, similar symptoms can be used as a basis to implement a series of diagnostic tests [65]. This method of raising clinical diagnostic standards was used in Wuhan, China.

In terms of health, we found that increasing the proportion of residents using at least basic sanitation services was significantly positive associated with the IR of COVID-19. For example, improving basic sanitation services and increasing contact between primary health workers and potential and diagnosed COVID-19 patients is very important. In particular, the government of Wuhan, China implemented nucleic acid testing on each resident via hospitals and primary health workers, thus enabling COVID-19 detection in a larger proportion and facilitating rapid control of COVID-19 risk transmission. Second, the numbers of hospital beds (per 1,000 people), nurses and midwives (per 1,000 people) were significantly negative associated with the IR and CMR of COVID-19, thus suggesting that COVID-19 risk should be controlled, and the number of hospital beds and nurses should be increased in a short period of time. The increase in the numbers of hospital beds and nurses can help achieve standardized management of patients and allow more medical resources to be concentrated on the treatment of severe cases. Some research has shown that some countries, such as Italy, China and the United States, have established Fangcang shelter hospitals or field hospitals and increased the numbers of regular hospital beds, intensive care beds and medical workers (by transferring resources from other regions and the military), reopened closed hospitals, and considered use of medical volunteers in the treatment of mild and severe COVID-19 cases; these measures are effective ways to reduce the IR and CMR [66, 67]. Third, among people infected with COVID-19, the proportion of the population spending more than 25% of household consumption or income on out-of-pocket health care expenses were significantly negative associated with the IR, whereas coverage with social insurance plans positively influences the IR. Higher income, enhanced health insurance coverage and decreased burden of medical treatment significantly increase the IR, thus suggesting that governments and health insurance providers should cooperate in the prevention and control of COVID-19. In addition to financial subsidies, the government should also reduce or grant exemptions for patient co-payments, to increase the possibility of COVID-19 patients receiving testing and treatment.

In science and technology, the number of researchers in R&D (per million people) was significantly positive associated with the DCI. This improvement includes facilitating health science and technology input, strengthening basic life science research, fostering international cooperation between science and technology (such as in the development and use of effective drugs), providing more convenient testing technology, shortening testing times, expanding the scale of detection, improving treatment technology and performing ongoing vaccine development to reduce present and future COVID-19 transmission. In addition, the poverty headcount ratio at the national poverty lines (percentage of population) was significantly negative associated with the IR, CMR and DCI. Increases in the population in poverty and in racial discrimination greatly diminish accessibility to medical services. Government and society must address these problems through economic stimulus plans, unemployment relief programs, welfare and health safeguarding measures, and plans to decrease health spending by these groups [68].

We also found that climatic factors (temperature, RH, precipitation and wind speed) were not significantly associated with COVID-19 risk, in agreement with the results of some studies [69]. However, previous studies have primarily considered the effects of single climate factors, thus potentially affecting the estimates of the results [70–72]. There is no sufficient evidence indicating that climate factors have specific effects on the spread of COVID-19. This study also shows that in the measurement of COVID-19 risk, the influences of other factors should be considered—such as the constraints of economic development, transportation and other factors—to improve understanding of the mechanisms underlying interrelationships among factors.

## 5. Limitations

This study has several limitations. First, we selected cross-sectional data for spatial analysis and regression modeling; therefore, the results may not reflect more changes in time, thus potentially decreasing the statistical ability to detect the relationships among various factors and COVID-19 risk. Second, owing to data matching across databases, some aspects of country/region data may have been lost, thus potentially affecting the spatial weight matrix estimation and regression modeling results. Third, because of the socio-ecological study design, we were unable to access data at the individual level, such as age, sex, occupation, economic and health status, and the actual exposure temperature of each person. However, future studies could adopt hybrid study designs, which use individual-level data from subpopulations to improve ecological extrapolation.

## 6. Conclusion

By using the data from 178 countries/regions, we found that socio-economic factors can significantly reduce the risk of COVID-19. As a next step in COVID-19 prevention, different countries/regions should focus on controlling urban populations, providing economic subsidies and medical resource supplies, and taking broad views of social welfare. Strategies may include population isolation, travel restrictions, case screening, cross-regional or national science and technology exchange to promote diagnosis and treatment, public welfare policy improvement, as well as decreasing the burden of low-income groups in obtaining medical treatment. Simultaneously, we must be alert to the COVID-19 risk in some countries in Africa and Asia, and must curb the second wave of COVID-19 transmission.

## Data Availability

The data used for the analyses are publicly available from the Johns Hopkins University Center for Systems Science and Engineering (https://github.com/CSSEGISandData/COVID-19), the World Bank (https://datacatalog.worldbank.org/dataset/world-development-indicators) and National Oceanic and Atmospheric Administration, Department of Commerce (https://catalog.data.gov/dataset/global-surface-summary-of-the-day-gsod).

https://datacatalog.worldbank.org/dataset/world-development-indicators

https://catalog.data.gov/dataset/global-surface-summary-of-the-day-gsod

## Acknowledgments

The authors would like to thank the National Natural Science Foundation of China and the National School of Development, Peking University, University of Michigan, and other members for their support and cooperation. We would also like to thank the study samples from The Center for Systems Science and Engineering, Johns Hopkins University (JHU-CSSE), Global Surface Summary of the Day (GSOD) from the Integrated Surface Hourly (ISH) dataset and World Development Indicators (WDI) dataset for providing the information in our research.

## Contributors

DS, XZ contributed to the conception and design of the project; DS, TZ, KH, XZ contributed to the analysis and interpretation of the data; MT, YZ contributed to the data acquisition and provided statistical analysis support; DS drafted the article. DS and XZ are the guarantors. The corresponding author attests that all listed authors meet authorship criteria and that no others meeting the criteria have been omitted.

## Patient and public involvement

This research was done without patient involvement. Patients were not invited to comment on the study design and were not consulted to develop patient relevant outcomes or interpret the results. Patients were not invited to contribute to the writing or editing of this document for readability or accuracy.

## Patient consent for publication

Not required.

## Ethics approval

This study was approved by the Ethics Committee of the Tongji Medical College, Huazhong University of Science and Technology (IORG No: IORG0003571).

---

**Table S1.**
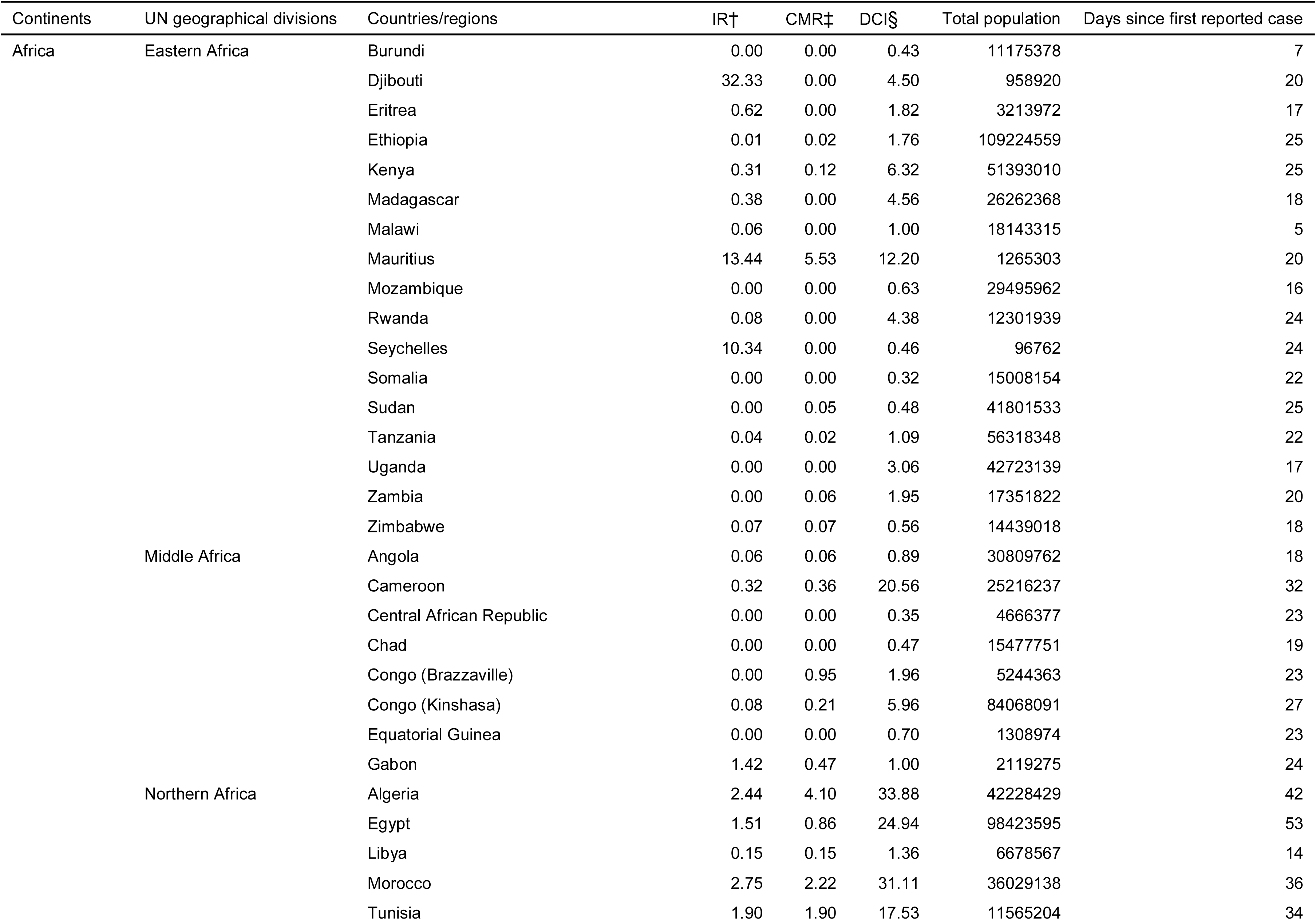
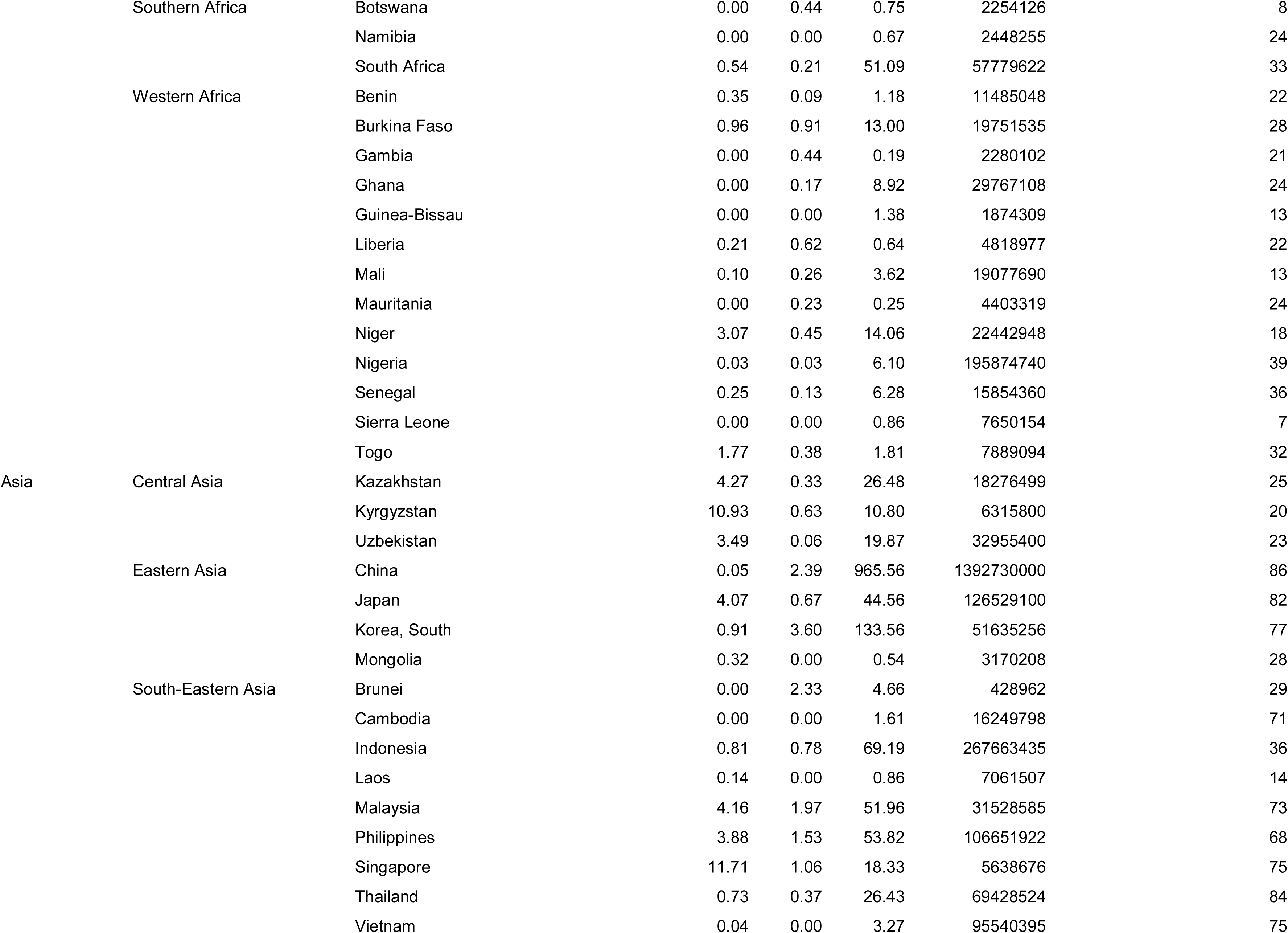
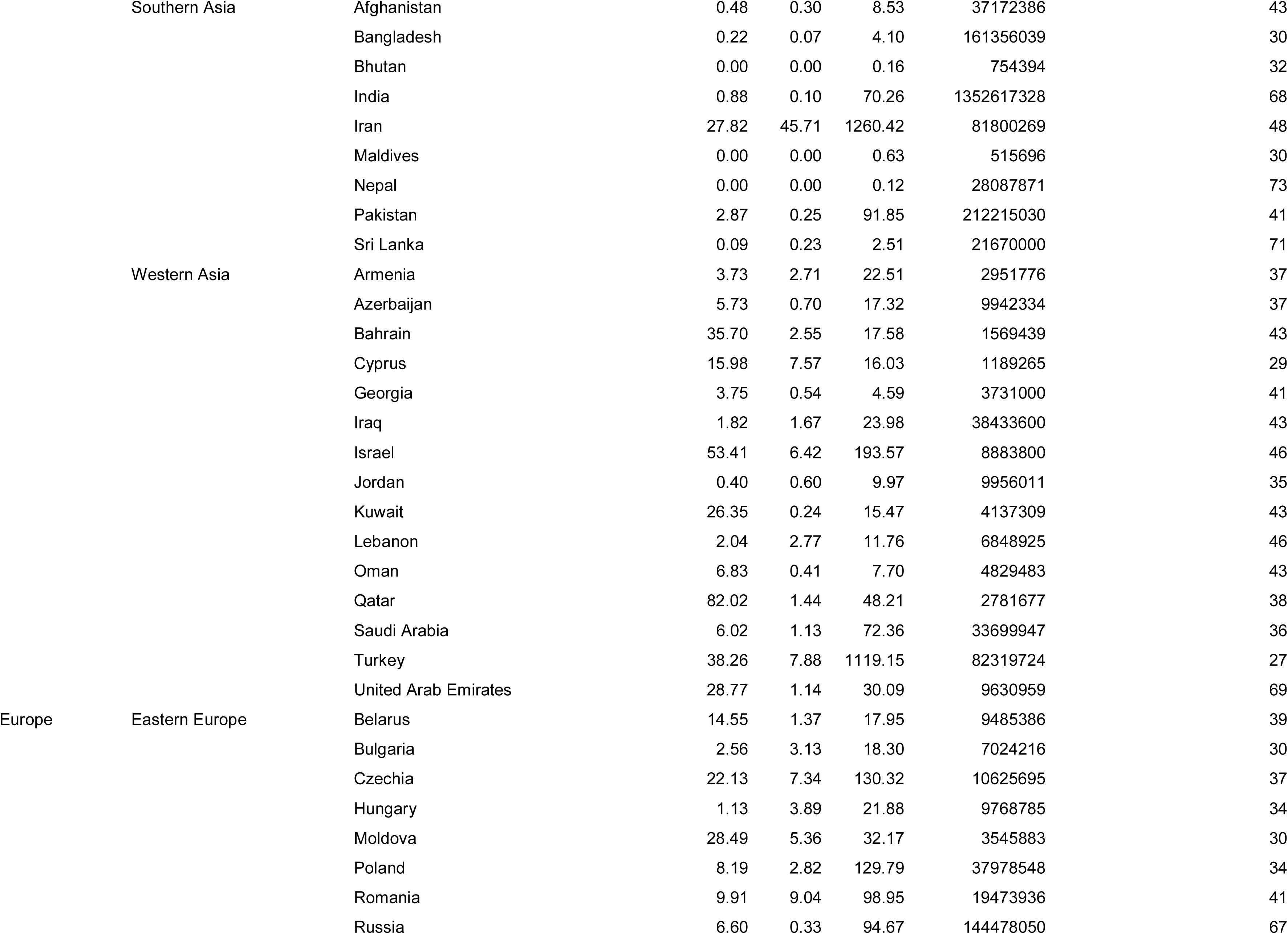
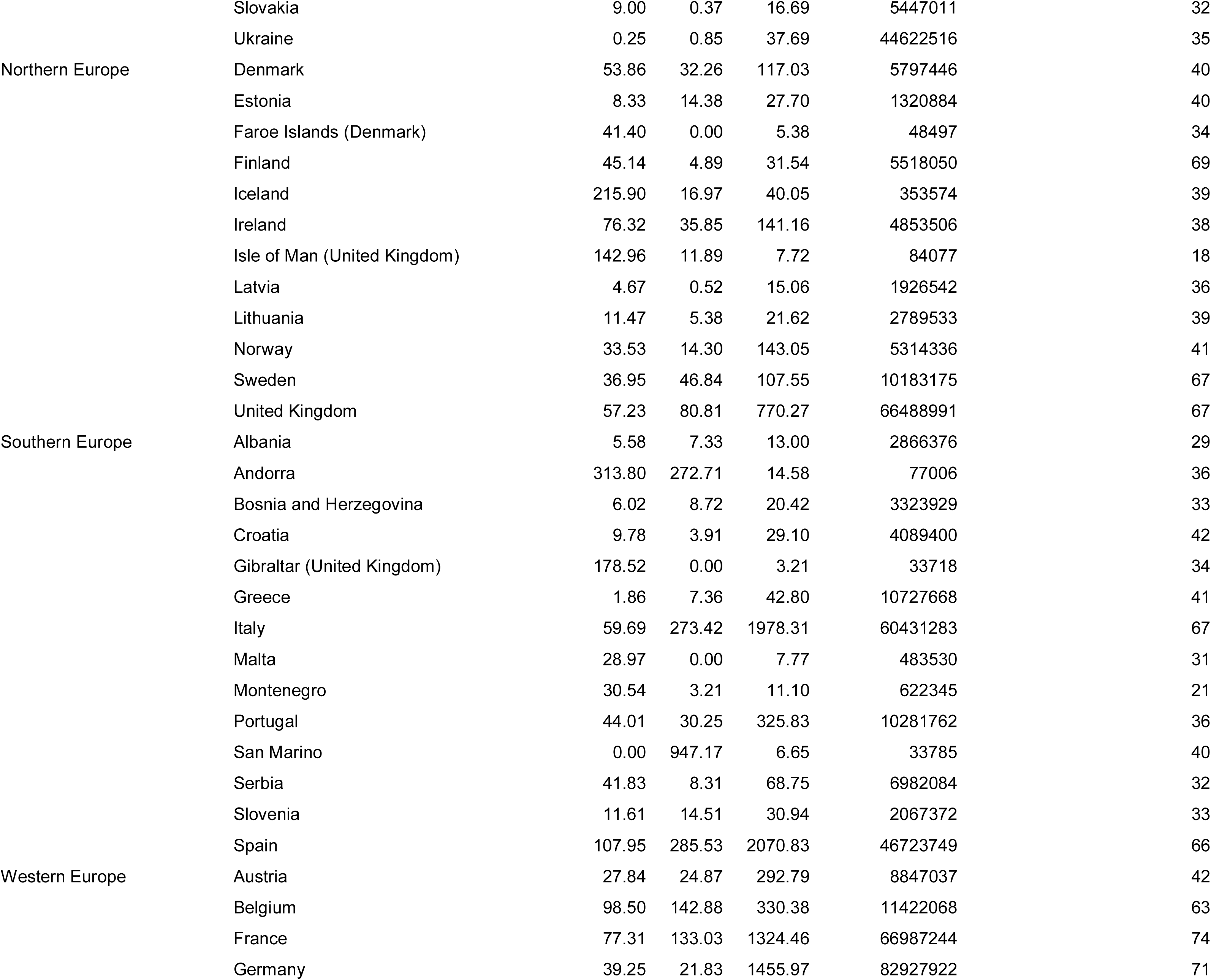
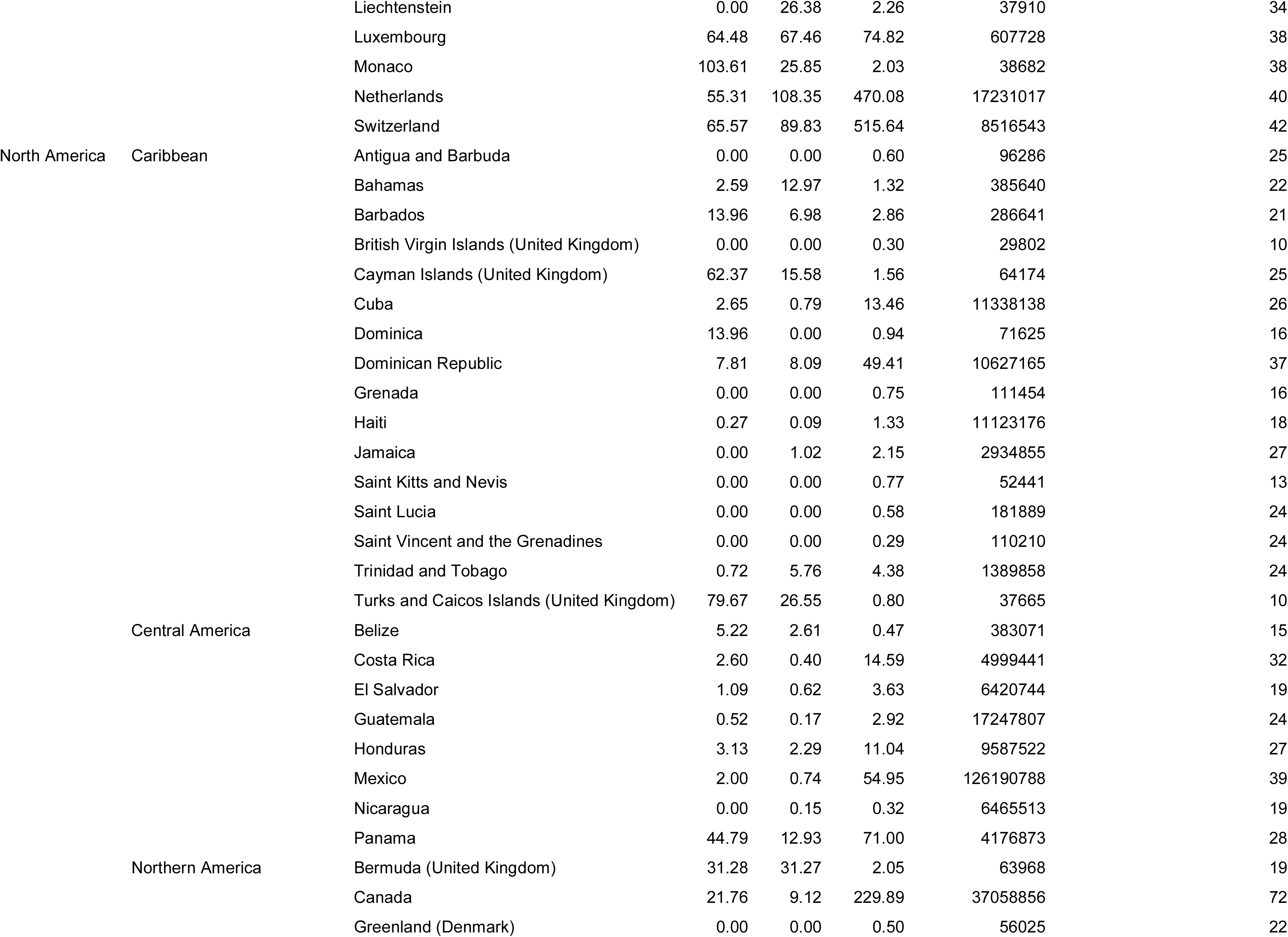
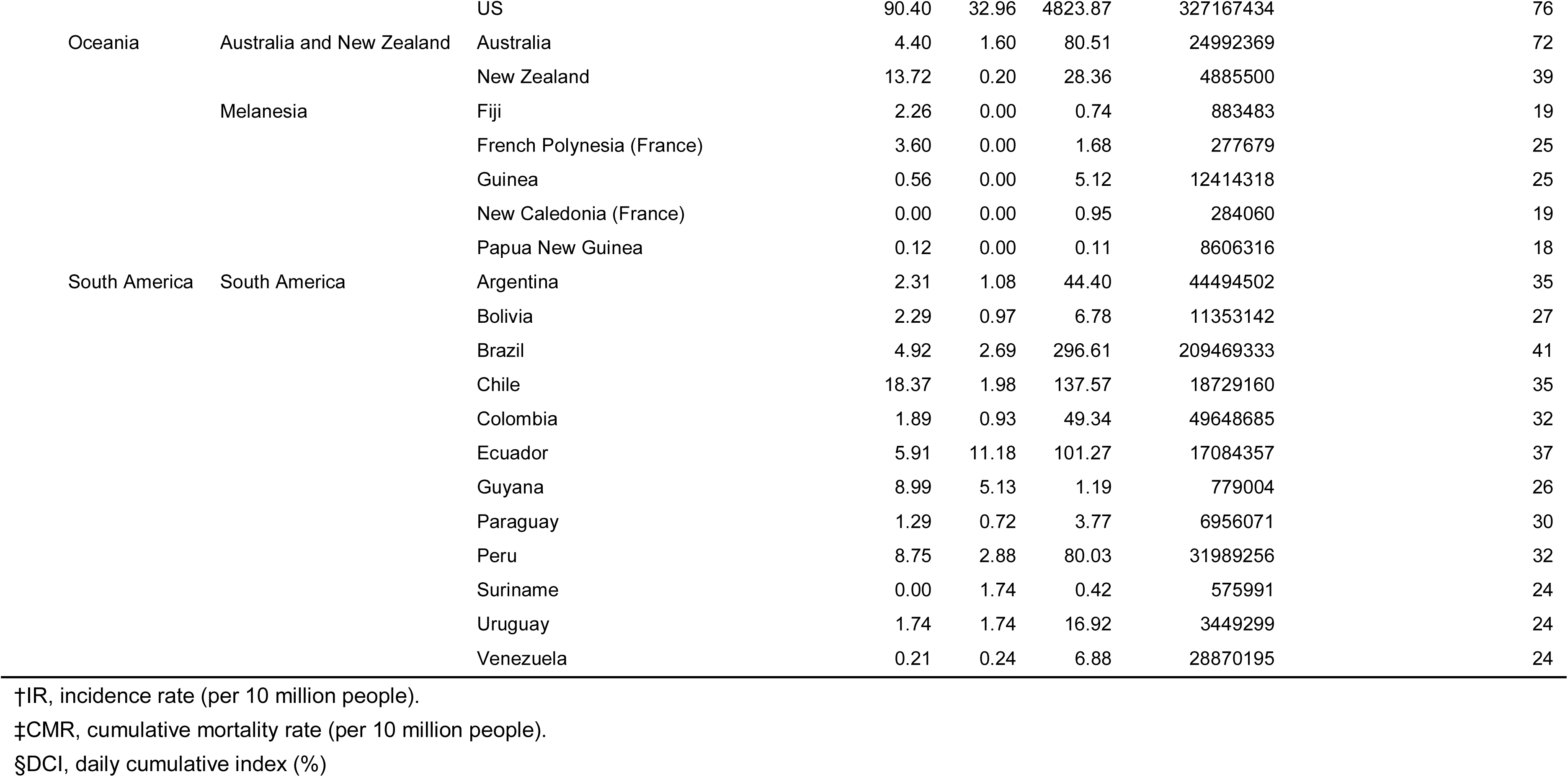
Characteristics of 178 countries/regions with reported cases of COVID-19 as of 6 April 2020

**Table S2.**
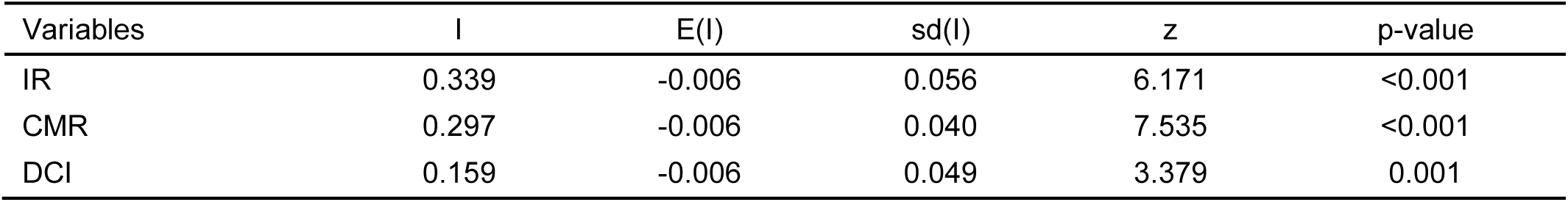
The Global Moran’s I index of IR, CMR and DCI for COVID-19

| Variables | I | E(I) | sd(I) | z | p-value |
| --- | --- | --- | --- | --- | --- |
| IR | 0.339 | -0.006 | 0.056 | 6.171 | <0.001 |
| CMR | 0.297 | -0.006 | 0.040 | 7.535 | <0.001 |
| DCI | 0.159 | -0.006 | 0.049 | 3.379 | 0.001 |

**Table S3.** Correlation matrix between socio-economic factors for the risk of COVID-19

| Variables | (1) | (2) | (3) | (4) | (5) | (6) | (7) | (8) | (9) | (10) | (11) | (12) | (13) | (14) | (15) | (16) | (17) | (18) | (19) | (20) | (21) | (22) | (23) | (24) | (25) | (26) | (27) | (28) |
| --- | --- | --- | --- | --- | --- | --- | --- | --- | --- | --- | --- | --- | --- | --- | --- | --- | --- | --- | --- | --- | --- | --- | --- | --- | --- | --- | --- | --- |
| (1) | 1.000 |  |  |  |  |  |  |  |  |  |  |  |  |  |  |  |  |  |  |  |  |  |  |  |  |  |  |  |
| (2) | -0.420* | 1.000 |  |  |  |  |  |  |  |  |  |  |  |  |  |  |  |  |  |  |  |  |  |  |  |  |  |  |
| (3) | 0.188* | -0.075 | 1.000 |  |  |  |  |  |  |  |  |  |  |  |  |  |  |  |  |  |  |  |  |  |  |  |  |  |
| (4) | 0.444* | -0.360* | 0.488* | 1.000 |  |  |  |  |  |  |  |  |  |  |  |  |  |  |  |  |  |  |  |  |  |  |  |  |
| (5) | 0.585* | -0.717* | 0.111 | 0.448* | 1.000 |  |  |  |  |  |  |  |  |  |  |  |  |  |  |  |  |  |  |  |  |  |  |  |
| (6) | 0.218* | -0.289* | -0.148* | 0.175* | 0.243* | 1.000 |  |  |  |  |  |  |  |  |  |  |  |  |  |  |  |  |  |  |  |  |  |  |
| (7) | 0.468* | -0.327* | 0.099 | 0.656* | 0.421* | 0.574* | 1.000 |  |  |  |  |  |  |  |  |  |  |  |  |  |  |  |  |  |  |  |  |  |
| (8) | -0.110 | -0.335* | -0.061 | -0.052 | -0.086 | 0.239* | 0.060 | 1.000 |  |  |  |  |  |  |  |  |  |  |  |  |  |  |  |  |  |  |  |  |
| (9) | -0.328* | 0.178* | -0.064 | -0.368* | -0.269* | -0.219* | -0.404* | -0.011 | 1.000 |  |  |  |  |  |  |  |  |  |  |  |  |  |  |  |  |  |  |  |
| (10) | 0.436* | -0.376* | 0.106 | 0.580* | 0.439* | 0.514* | 0.890* | 0.013 | -0.194* | 1.000 |  |  |  |  |  |  |  |  |  |  |  |  |  |  |  |  |  |  |
| (11) | 0.376* | -0.602* | 0.293* | 0.407* | 0.497* | 0.195* | 0.304* | 0.380* | -0.296* | 0.268* | 1.000 |  |  |  |  |  |  |  |  |  |  |  |  |  |  |  |  |  |
| (12) | 0.573* | -0.653* | 0.106 | 0.559* | 0.858* | 0.345* | 0.557* | -0.158* | -0.383* | 0.536* | 0.454* | 1.000 |  |  |  |  |  |  |  |  |  |  |  |  |  |  |  |  |
| (13) | -0.503* | 0.632* | -0.085 | -0.367* | -0.858* | -0.110 | -0.351* | 0.106 | 0.283* | -0.360* | -0.462* | -0.807* | 1.000 |  |  |  |  |  |  |  |  |  |  |  |  |  |  |  |
| (14) | -0.521* | 0.690* | -0.116 | -0.455* | -0.861* | -0.302* | -0.457* | 0.107 | 0.381* | -0.455* | -0.534* | -0.913* | 0.858* | 1.000 |  |  |  |  |  |  |  |  |  |  |  |  |  |  |
| (15) | 0.217* | -0.033 | 0.006 | 0.228* | 0.164* | 0.397* | 0.426* | 0.050 | -0.353* | 0.279* | 0.163* | 0.193* | -0.120 | -0.191* | 1.000 |  |  |  |  |  |  |  |  |  |  |  |  |  |
| (16) | 0.503* | -0.493* | 0.288* | 0.659* | 0.567* | 0.416* | 0.758* | 0.182* | -0.419* | 0.649* | 0.613* | 0.616* | -0.507* | -0.600* | 0.285* | 1.000 |  |  |  |  |  |  |  |  |  |  |  |  |
| (17) | 0.534* | -0.674* | 0.088 | 0.405* | 0.875* | 0.289* | 0.401* | -0.063 | -0.221* | 0.412* | 0.451* | 0.837* | -0.825* | -0.832* | 0.132 | 0.568* | 1.000 |  |  |  |  |  |  |  |  |  |  |  |
| (18) | 0.556* | -0.662* | 0.218* | 0.497* | 0.660* | 0.441* | 0.571* | 0.276* | -0.339* | 0.525* | 0.600* | 0.676* | -0.594* | -0.690* | 0.190* | 0.747* | 0.658* | 1.000 |  |  |  |  |  |  |  |  |  |  |
| (19) | 0.388* | -0.715* | -0.059 | 0.358* | 0.607* | 0.546* | 0.594* | 0.524* | -0.340* | 0.540* | 0.587* | 0.702* | -0.558* | -0.662* | 0.219* | 0.650* | 0.606* | 0.728* | 1.000 |  |  |  |  |  |  |  |  |  |
| (20) | -0.274* | 0.931* | -0.038 | -0.249* | -0.606* | -0.246* | -0.230* | -0.427* | 0.133 | -0.272* | -0.544* | -0.561* | 0.564* | 0.613* | 0.055 | -0.390* | -0.594* | -0.569* | -0.703* | 1.000 |  |  |  |  |  |  |  |  |
| (21) | -0.018 | -0.069 | 0.028 | -0.088 | 0.026 | 0.273* | -0.071 | 0.090 | 0.335* | 0.045 | 0.016 | 0.026 | 0.065 | 0.021 | -0.177* | -0.063 | 0.073 | 0.045 | 0.067 | -0.093 | 1.000 |  |  |  |  |  |  |  |
| (22) | -0.105 | 0.056 | 0.003 | -0.110 | -0.103 | 0.235* | -0.097 | 0.050 | 0.280* | -0.013 | -0.105 | -0.105 | 0.193* | 0.160* | -0.174* | -0.135 | -0.061 | -0.066 | -0.067 | 0.011 | 0.883* | 1.000 |  |  |  |  |  |  |
| (23) | -0.065 | 0.020 | -0.006 | -0.051 | -0.026 | -0.078 | -0.031 | 0.019 | 0.147* | -0.038 | -0.010 | -0.020 | -0.011 | 0.010 | -0.347* | -0.027 | 0.004 | -0.026 | 0.008 | -0.050 | 0.133 | 0.116 | 1.000 |  |  |  |  |  |
| (24) | -0.355* | 0.453* | 0.027 | -0.237* | -0.639* | -0.222* | -0.341* | 0.045 | 0.189* | -0.312* | -0.371* | -0.676* | 0.658* | 0.660* | -0.111 | -0.456* | -0.727* | -0.500* | -0.540* | 0.447* | -0.080 | 0.018 | -0.101 | 1.000 |  |  |  |  |
| (25) | 0.459* | -0.399* | 0.040 | 0.509* | 0.447* | 0.486* | 0.778* | 0.217* | -0.375* | 0.666* | 0.419* | 0.584* | -0.392* | -0.501* | 0.275* | 0.724* | 0.449* | 0.600* | 0.728* | -0.329* | -0.029 | -0.108 | -0.004 | -0.439* | 1.000 |  |  |  |
| (26) | 0.368* | -0.324* | -0.020 | 0.542* | 0.381* | 0.509* | 0.828* | 0.200* | -0.399* | 0.703* | 0.341* | 0.533* | -0.339* | -0.440* | 0.281* | 0.730* | 0.373* | 0.568* | 0.667* | -0.260* | -0.076 | -0.108 | -0.009 | -0.362* | 0.838* | 1.000 |  |  |
| (27) | 0.498* | -0.598* | 0.033 | 0.368* | 0.586* | 0.451* | 0.544* | 0.350* | -0.285* | 0.482* | 0.551* | 0.630* | -0.568* | -0.640* | 0.249* | 0.704* | 0.664* | 0.697* | 0.782* | -0.534* | 0.089 | -0.055 | 0.036 | -0.586* | 0.693* | 0.617* | 1.000 |  |
| (28) | -0.019 | -0.031 | -0.052 | -0.203* | 0.013 | 0.019 | -0.175* | 0.053 | 0.141 | -0.113 | -0.039 | -0.118 | -0.015 | 0.049 | -0.013 | -0.144 | -0.096 | -0.114 | -0.068 | -0.041 | 0.050 | -0.004 | -0.067 | 0.167* | -0.183* | -0.167* | -0.110 | 1.000 |

\* shows significance at the .05 level

**Table S4.** The sensitive analysis of single-factor negative binominal regression for the risk of COVID-19

| Indicators | IR |  |  |  | CMR |  |  |  | DCI |  |  |  |
| --- | --- | --- | --- | --- | --- | --- | --- | --- | --- | --- | --- | --- |
|  | IRR† | 95%CI‡<br>(Lower) | 95%CI<br>(Upper) | p-value | IRR | 95%CI<br>(Lower) | 95%CI<br>(Upper) | p-value | IRR | 95%CI<br>(Lower) | 95%CI<br>(Upper) | p-value |
| Max temperature (Celsius) | 0.911 | 0.881 | 0.942 | <0.001 | 0.960 | 0.860 | 1.072 | 0.470 | 0.838 | 0.801 | 0.877 | <0.001 |
| Min temperature (Celsius) | 0.939 | 0.908 | 0.971 | <0.001 | 0.983 | 0.900 | 1.075 | 0.713 | 0.845 | 0.812 | 0.880 | <0.001 |

†IRR is short for incidence rate ratio.
‡CI is short for confidence interval.

**Table S5.**
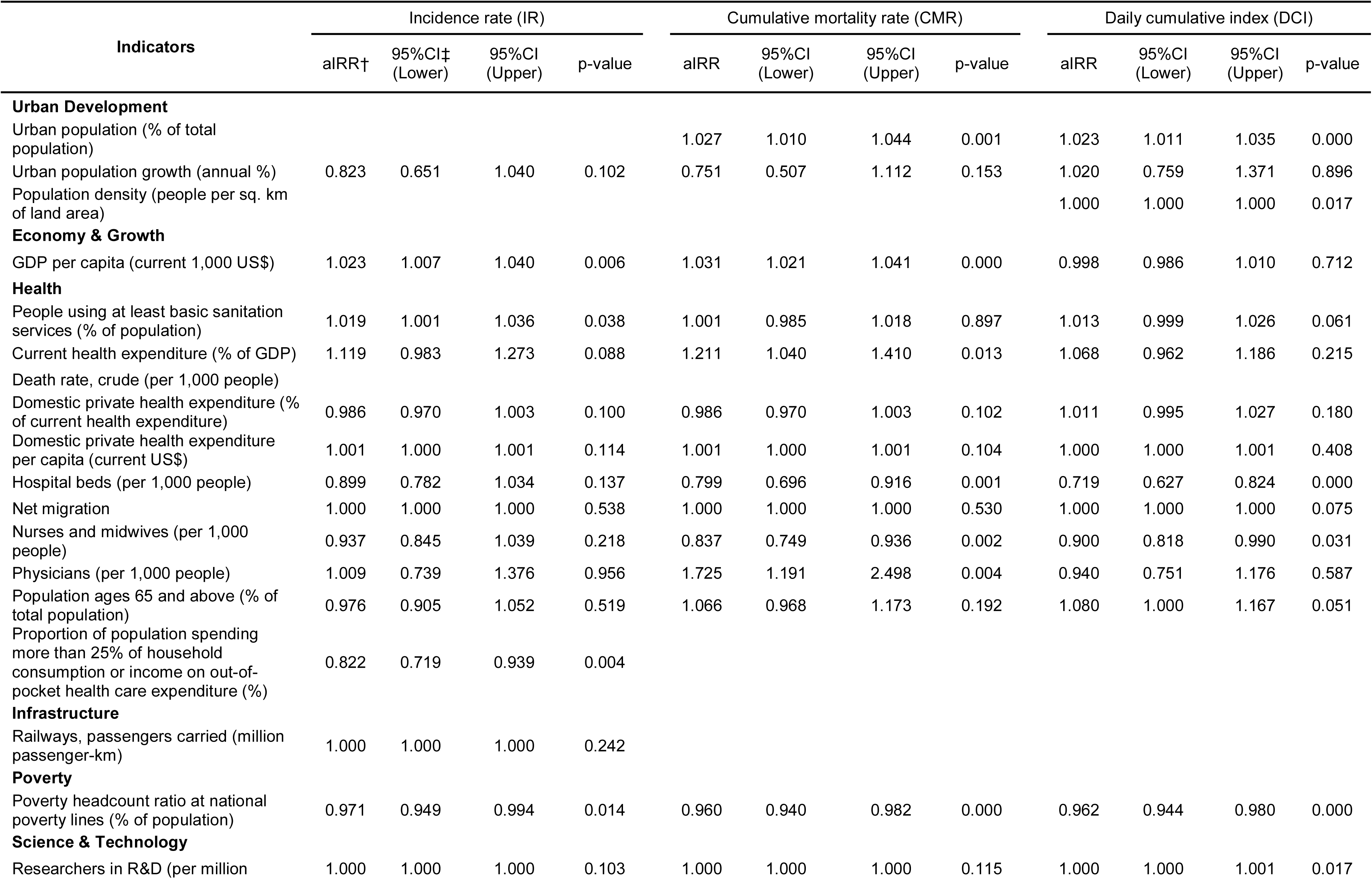
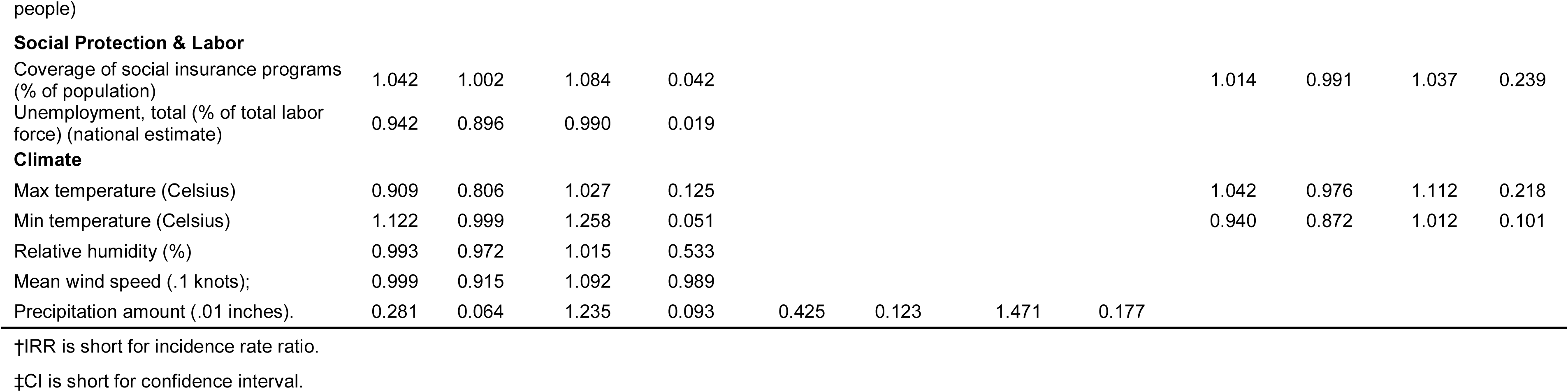
The sensitive analysis of multiple-factor negative binominal regression for the risk of COVID-19

| Indicators | Incidence rate (IR) |  |  |  | Cumulative mortality rate (CMR) |  |  |  | Daily cumulative index (DCI) |  |  |  |
| --- | --- | --- | --- | --- | --- | --- | --- | --- | --- | --- | --- | --- |
|  | aIRR† | 95%CI‡<br>(Lower) | 95%CI<br>(Upper) | p-value | aIRR | 95%CI<br>(Lower) | 95%CI<br>(Upper) | p-value | aIRR | 95%CI<br>(Lower) | 95%CI<br>(Upper) | p-value |
| **Urban Development** |  |  |  |  |  |  |  |  |  |  |  |  |
| Urban population (% of total population) |  |  |  |  | 1.027 | 1.010 | 1.044 | 0.001 | 1.023 | 1.011 | 1.035 | 0.000 |
| Urban population growth (annual %) | 0.823 | 0.651 | 1.040 | 0.102 | 0.751 | 0.507 | 1.112 | 0.153 | 1.020 | 0.759 | 1.371 | 0.896 |
| Population density (people per sq. km of land area) |  |  |  |  |  |  |  |  | 1.000 | 1.000 | 1.000 | 0.017 |
| **Economy & Growth** |  |  |  |  |  |  |  |  |  |  |  |  |
| GDP per capita (current 1,000 US\$) | 1.023 | 1.007 | 1.040 | 0.006 | 1.031 | 1.021 | 1.041 | 0.000 | 0.998 | 0.986 | 1.010 | 0.712 |
| **Health** |  |  |  |  |  |  |  |  |  |  |  |  |
| People using at least basic sanitation services (% of population) | 1.019 | 1.001 | 1.036 | 0.038 | 1.001 | 0.985 | 1.018 | 0.897 | 1.013 | 0.999 | 1.026 | 0.061 |
| Current health expenditure (% of GDP) | 1.119 | 0.983 | 1.273 | 0.088 | 1.211 | 1.040 | 1.410 | 0.013 | 1.068 | 0.962 | 1.186 | 0.215 |
| Death rate, crude (per 1,000 people) |  |  |  |  |  |  |  |  |  |  |  |  |
| Domestic private health expenditure (% of current health expenditure) | 0.986 | 0.970 | 1.003 | 0.100 | 0.986 | 0.970 | 1.003 | 0.102 | 1.011 | 0.995 | 1.027 | 0.180 |
| Domestic private health expenditure per capita (current US\$) | 1.001 | 1.000 | 1.001 | 0.114 | 1.001 | 1.000 | 1.001 | 0.104 | 1.000 | 1.000 | 1.001 | 0.408 |
| Hospital beds (per 1,000 people) | 0.899 | 0.782 | 1.034 | 0.137 | 0.799 | 0.696 | 0.916 | 0.001 | 0.719 | 0.627 | 0.824 | 0.000 |
| Net migration | 1.000 | 1.000 | 1.000 | 0.538 | 1.000 | 1.000 | 1.000 | 0.530 | 1.000 | 1.000 | 1.000 | 0.075 |
| Nurses and midwives (per 1,000 people) | 0.937 | 0.845 | 1.039 | 0.218 | 0.837 | 0.749 | 0.936 | 0.002 | 0.900 | 0.818 | 0.990 | 0.031 |
| Physicians (per 1,000 people) | 1.009 | 0.739 | 1.376 | 0.956 | 1.725 | 1.191 | 2.498 | 0.004 | 0.940 | 0.751 | 1.176 | 0.587 |
| Population ages 65 and above (% of total population) | 0.976 | 0.905 | 1.052 | 0.519 | 1.066 | 0.968 | 1.173 | 0.192 | 1.080 | 1.000 | 1.167 | 0.051 |
| Proportion of population spending more than 25% of household consumption or income on out-of-pocket health care expenditure (%) | 0.822 | 0.719 | 0.939 | 0.004 |  |  |  |  |  |  |  |  |
| **Infrastructure** |  |  |  |  |  |  |  |  |  |  |  |  |
| Railways, passengers carried (million passenger-km) | 1.000 | 1.000 | 1.000 | 0.242 |  |  |  |  |  |  |  |  |
| **Poverty** |  |  |  |  |  |  |  |  |  |  |  |  |
| Poverty headcount ratio at national poverty lines (% of population) | 0.971 | 0.949 | 0.994 | 0.014 | 0.960 | 0.940 | 0.982 | 0.000 | 0.962 | 0.944 | 0.980 | 0.000 |
| **Science & Technology** |  |  |  |  |  |  |  |  |  |  |  |  |
| Researchers in R&D (per million) | 1.000 | 1.000 | 1.000 | 0.103 | 1.000 | 1.000 | 1.000 | 0.115 | 1.000 | 1.000 | 1.001 | 0.017 |
| **Social Protection & Labor** |  |  |  |  |  |  |  |  |  |  |  |  |
| Coverage of social insurance programs<br>(% of population) | 1.042 | 1.002 | 1.084 | 0.042 |  |  |  |  | 1.014 | 0.991 | 1.037 | 0.239 |
| Unemployment, total (% of total labor<br>force) (national estimate) | 0.942 | 0.896 | 0.990 | 0.019 |  |  |  |  |  |  |  |  |
| **Climate** |  |  |  |  |  |  |  |  |  |  |  |  |
| Max temperature (Celsius) | 0.909 | 0.806 | 1.027 | 0.125 |  |  |  |  | 1.042 | 0.976 | 1.112 | 0.218 |
| Min temperature (Celsius) | 1.122 | 0.999 | 1.258 | 0.051 |  |  |  |  | 0.940 | 0.872 | 1.012 | 0.101 |
| Relative humidity (%) | 0.993 | 0.972 | 1.015 | 0.533 |  |  |  |  |  |  |  |  |
| Mean wind speed (.1 knots); | 0.999 | 0.915 | 1.092 | 0.989 |  |  |  |  |  |  |  |  |
| Precipitation amount (.01 inches). | 0.281 | 0.064 | 1.235 | 0.093 | 0.425 | 0.123 | 1.471 | 0.177 |  |  |  |  |

†IRR is short for incidence rate ratio.
‡CI is short for confidence interval.

